# Voluntary Cough Acoustic Signals Differentiate Disease State and Swallowing Safety Status: A Data Set in Healthy Controls vs. Motor Speech Disordered Patients

**DOI:** 10.1101/2025.09.10.25335329

**Authors:** Stephanie Watts, Shaheen N. Awan, Mohamed Ebraheem, Elijah Moothedan, Teressa Pitts, Yael Bensoussan

**Affiliations:** Department of Otolaryngology Head and Neck Surgery, College of Medicine, University of South Florida; School of Communication Sciences and Disorders & The Communication Technologies Research Center, University of Central Florida; Charles E. Schmidt College of Medicine, Florida Atlantic University; Department of Health Sciences, University of Missouri

**Keywords:** Dysphagia, MFCC analysis, Amyotrophic Lateral Sclerosis, Parkinson’s disease, Dystussia, Acoustic analysis

## Abstract

**Objective:** Changes in voluntary cough airflow are an established detector of swallowing safety in neurogenic dysphagia. The objective of the current project was to determine if acoustic measures of voluntary cough acoustic signals could be used to differentiate (1) healthy versus diseased state, (2) between disordered groups with Parkinson’s disease (PD) and Amyotrophic Lateral Sclerosis (ALS) and (3) those who have airway invasion during swallowing and those who do not (safe vs. unsafe swallows).

**Methods:** Videofluoroscopic swallow studies (VFSS) and voluntary coughs were obtained from PD and ALS participants (n=10 and n=13 respectively) and age matched controls (n=10). Participants with PD and ALS were striated into safe and unsafe swallowing groups using the penetration aspiration scale (PAS) on VFSS. Cough acoustic waveforms were processed with Python 3.11.4 and audio signal processing library Librosa 0.10.0 for extracting Mel Frequency Cepstrum Coefficients (MFCCs) and related acoustic measures of zero crossing rate (ZCR), spectral centroid, spectral bandwidth, spectral roll off, and spectral contrast.

**Results:** MFCC 5 emerged as the strongest acoustic discriminator between healthy and disordered populations with a receiver operating characteristic (ROC) area under the (AUC) = 0.80, sensitivity = 0.83, specificity = 0.73. MFCC 5 was also observed to successfully categorize healthy vs. PD subjects (ROC AUC = 0.84), while ZCR successfully categorized healthy vs. ALS subjects (AUC = 0.79). MFCC 2 successfully categorized ALS vs. PD subjects (AUC = 0.79), and MFCC 13 emerged as the best acoustic classifier of subjects with safe vs. unsafe swallow (AUC = 0.81).

**Conclusions:** Cough creates a distinct sound pattern during the transition from a compression phase, or closure of the glottis, to expiration. These results demonstrate that, even with a conservative approach to acoustic analysis of voluntary cough signals, MFCC analyses, along with other acoustic features including ZCR, show the potential for distinguishing between disease states and swallowing safety status with strong classification accuracy. Further research on the physiologic etiology of these acoustic differences is needed to provide thorough interpretation of the characteristics of healthy vs. disordered coughs.

## Introduction

Amyotrophic Lateral Sclerosis (ALS) and Parkinson’s disease (PD) are progressive and debilitating neurodegenerative diseases. Dysphagia, or swallowing impairment, is prevalent in both patient populations ^1–2^ with dystussia (pervasive respiratory and cough impairment)contributing to dysphagia ^3–5^. Presenting symptoms and disease onset are heterogeneous; thus, dysphagia symptoms manifest differently across patients and can be difficult to assess clinically. Individuals with ALS may present with bulbar onset of the disease in which case lingual and masticatory deficits are prominent features of dysphagia. In the spinal onset variant of ALS, limb motor impairment is the first salient feature identified ^6^. Patients with PD may also present with a variety of dysphagia symptoms over the course of the disease ^7^. Although the exact pathophysiology of dysphagia in PD is unknown, patients may present with dysphagia preclinically in the prodromal phase ^1,8^.

Despite onset type, patients with ALS are likely to develop impaired reflexive cough ^6^ . In ALS, neurodegeneration of respiratory motor neuron pools leads to impairments in neural activation and muscle coordination, weakness and atrophy of respiratory muscles, and weakened airway defenses, with combined neural, airway, pulmonary, and neuromuscular changes resulting in deterioration of respiratory-related functions including cough and swallowing ^9^. Work by Plowman and colleagues (2016) has shown that impaired voluntary cough function identifies patients with ALS who are at risk for penetration/aspiration. The authors postulated that there might be clinical utility for the use of voluntary cough airflow testing to detect presence of dysphagia in this patient population. Cough dysfunction has been reported in more than 50% of asymptomatic PD patients ^10^ with the motor component of cough primarily impaired in the early stages of the disease, and both the motor and sensory components of cough may be impaired in advanced stages of the disease ^11^. Because of cough reflex impairment and chest wall rigidity, these impairments may contribute to silent aspiration and increased risk of pneumonia ^11–12^. Patients with PD develop chest wall rigidity and subsequent respiratory dysfunction ^13^. Literature suggests also that voluntary cough airflow may help differentiate safe and unsafe swallowers ^3–4^. Although cough may be a clinically useful tool to determine compromised swallowing function or the ability to clear aspirant material, cough evaluation methodology and implementation is widely varied throughout the literature ^14^.

Methodology for common clinical cough testing includes subjective assessments of strength and/or weakness ^14^. This is regardless of clinician demographics such as years of experience, certification status, and/or practice setting. Unfortunately, it has been shown that subjective perceptual assessments of cough are unreliable and are not able to reliably detect swallowing impairment. For example, McCullough et al. (2005) reported high sensitivities (<79%) but low specificities (<42%) for detection of aspiration using perceptual ratings. The authors reported that almost half of the at-risk patients were wrongly categorized based on a “weak” cough rating ^15^. As an additional or possibly alternative method to perceptual ratings, acoustic measurements of voluntary cough sounds have been investigated in healthy adults. Olia et al. (2000) studied 234 cough patterns in healthy male and female adults. The authors subdivided cough patterns into distinct anthropomorphic features consisting of three components 1) explosive phase (phase timing and amplitude, 2) continuous phase (phase timing), and 3) the variable phase (timing). Authors reported that there were significant differences found between sex for the length of the expulsive phase, frequency of the first phase, and highest continuous frequency of the continuous phase. Voluntary cough sound patterns have been shown to detect abnormality in pulmonary lung function in human models. The focus of other studies investigating acoustic cough signals in healthy adults have been, length of cough segments and spectral frequency analysis^17^ as well as measures including sound power/energy, signal rise time, and duration ^18–19^.

### Cough Analyses Using Mel-Frequency Cepstral Coefficients (MFCCs)

A promising form of spectral analysis that has been used in various cough identification and classification studies has been the use of mel-frequency cepstral coefficients (MFCC’s). MFCCs provide a comprehensive representation of an audio signal via a collective set of features which describe the overall shape of the spectral envelope. MFCCs have been used in the identification and counting of cough vs. non-cough sounds ^20–23^, as well as the categorization of cough signals in terms of underlying disorder. As examples, several studies have successfully used MFCCs in the classification of Covid vs. healthy cough, Covid vs. asthmatic cough, and in the classification of wet vs. dry cough signals ^24–27^.

MFCCs were originally developed for speech recognition and have found diverse use as voice descriptors, for example in emotions recognition or speech disorder classification ^28^. The MFCC work by partitioning the speech frequency into overlapping mel-frequency filter banks, followed by the application of cepstral and cosine transformations on each bank ^29^. MFCC’s are then obtained from the short-term power spectrum, based on a linear cosine transform of the log power spectrum on a nonlinear Mel scale. MFCCs have been used in voice recognition applications and have also been used to describe the quality or “timbre” of both musical and voice signals as well as vocal tract characteristics^30–31^. Since MFCC analysis does not require periodicity in the acoustic signal or any form of pitch detection, the method is applicable to the cough signal ^32^.

### Study Purpose

While the underlying physiology of cough may be associated with certain perceptual characteristics, assessment of strength and effectiveness of cough via perceptual rating scales has been reported to be inconsistent^33^. Although there are limitations in acoustic cough analysis, objective methods are necessary to provide objective qualification and quantification of a subjective physiologic events such as cough^34^. Objective quantification of cough may help to determine differences in airway protection between healthy vs. disordered individuals, change in airway protection in specific diseases over time, or monitor improvement in airway protection following therapeutic intervention^35^. While MFCCs have been used in the classification of motor speech disordered voice signals in PD and ALS^36–37, 32,29,38–39,28,40^ we are unaware of studies that have used MFCC’s and related acoustic features to not only distinguish healthy vs. motor speech disordered cough signals, but also attempt to classify within motor speech disorders (PD vs. ALS) and to classify safe vs. unsafe swallows within a motor speech disordered cohort.

The following research questions were addressed in this study:

1. Can acoustic features of voluntary cough differentiate healthy vs neurologically disordered subjects (subjects with Parkinson’s disease [PD] and Amyotrophic Lateral Sclerosis [ALS])?
2. Can acoustic features of voluntary cough differentiate between neurologically-based disorders (PD vs. ALS)?
3. In motor speech disordered subjects (PD and ALS), can acoustic features of voluntary cough differentiate between safe vs. unsafe swallow?

## Methods

This study received approval by the University of South Florida Institutional Review Board (#00023151).

### Subjects

Participants included 33 adult participants composed of 10 healthy age matched controls with no known history of pulmonary disease (5 male and 5 female; average age (yrs) = 59, range = 42-75); 13 patients diagnosed with ALS according the El Escorial Criteria (Brooks et al., 2000) with bulbar, spinal, and mixed disease onsets (5 male, 8 female; average age (yrs) = 62.4, range = 39-73); and 10 patients diagnosed with PD (8 male, 2 female; average age (yrs) = 69, range = 58-81). Healthy controls were recruited through a convenience sample from the local community and university staff. All patients were recruited from the Morsani Medical Center’s neurology clinics. Participants with Parkinson’s disease (PD) or Amyotrophic Lateral Sclerosis (ALS) were diagnosed by fellowship-trained neurologists specializing in movement disorders. ALS diagnosis was confirmed according to El Escorial criteria and PD staging provided by Hoehn & Yahr scale, ensuring that participants met the established clinical and diagnostic standards for their respective conditions. Mean age of the sample was 65.2 years (range 39-81 years). All subjects scored > 26 on the MMSE and were deemed appropriate to participate in the cough and swallow evaluation protocols. Table 1 summarizes the demographic and disease data for each subject within the ALS cohort and Table 2 summarizes the demographic and disease data for each subject within the PD cohort. In addition to the diagnoses above, specific inclusion criteria included: 1) cognition within normal limits as determined by >24 points on the Mini Mental Status Exam (Reisberg et al., 1982). Specific exclusion criteria include: 1) presence of tracheotomy or mechanical ventilation; 2) presence of diaphragmatic pacer; 3) diagnosis of significant concurrent respiratory disease (e.g., COPD); or 4) allergies to barium.

**Table 1.**
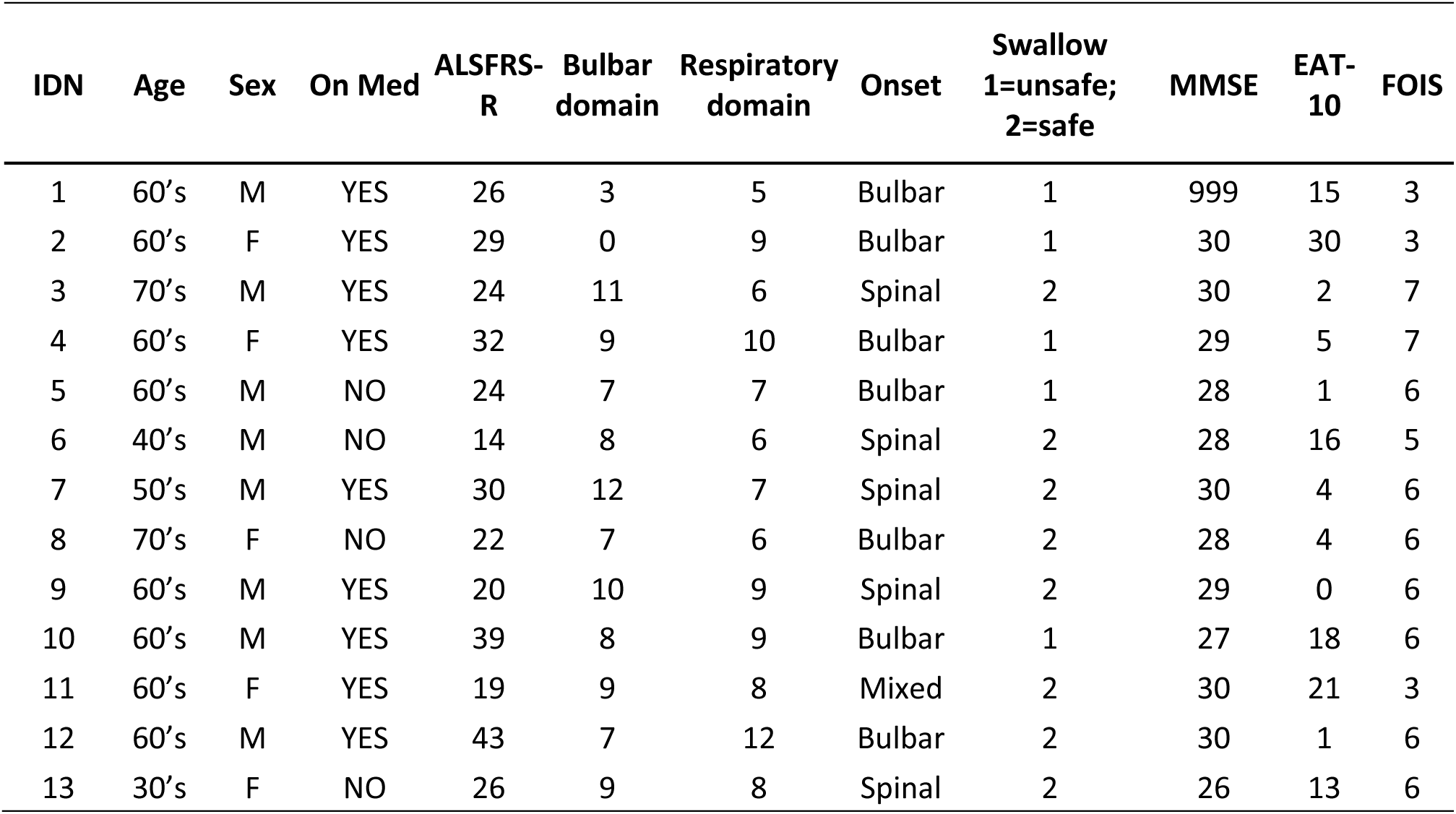
Demographic Information for the ALS disease cohort including age, gender, medication status for on Riluzole/Neudexta, ALS Functional Rating Scale scores (ALSFRS) including both bulbar and respiratory domains, swallowing safety group with PAS ≤ 2 being safe, mini mental status examination (MMSE), Eating assessment tool (EAT-10) and Functional Oral Intake Scale (FOIS).

**Table 2.** Demographic Information for the PD disease cohort including age, sex, medication status for Levodopa/Caridopa, Hoehn & Yahr scores, swallowing safety group with PAS score ≤ 2 being safe, MMSE, EAT-10 and FOIS.

| IDN | Age | Sex | On Med | Hoehn & Yahr | Swallow 1-unsafe;<br>2=safe | MMSE | EAT-10 | FOIS |
| --- | --- | --- | --- | --- | --- | --- | --- | --- |
| 1 | 70's | M | YES | 3 | 1 | 29 | 11 | 7 |
| 2 | 70's | M | YES | 3 | 2 | 30 | 28 | 5 |
| 3 | 60's | M | NO | 3 | 2 | 29 | 8 | 7 |
| 4 | 50's | F | YES | n/a | 2 | 29 | 24 | 6 |
| 5 | 80's | M | YES | 4 | 2 | 24 | 7 | n/a |
| 6 | 50's | M | YES | 3 | 2 | 30 | 2 | 7 |
| 7 | 80's | M | YES | 2 | 2 | 30 | 0 | 7 |
| 8 | 60's | M | YES | 3 | 1 | 29 | 7 | 6 |
| 9 | 60's | F | YES | n/a | 2 | 30 | n/a | n/a |
| 10 | 70's | M | YES | n/a | 1 | 30 | 15 | 5 |

Note: PD = Parkinson's disease, EAT-10 = Eating assessment tool, PAS = Penetration Aspiration Scale, MMSE = mini mental status examination, FOIS = Functional Oral Intake Scale

### Testing Protocol

The acoustic data analyzed in this study was collected as part of a larger set of evaluation testing that included swallowing, respiratory function, and voluntary cough testing (the focus of this study). Once enrolled, participants were assigned a study number and underwent both evaluation of swallowing, respiratory function, and voluntary acoustic cough testing. The swallowing study and respiratory acoustic testing procedures were performed on the same day. Data collection was counterbalanced to account for fatigue in these patient populations. Assessments included: 1) Mini Mental Status Examination (MMSE) to assess for ability to complete directions for voluntary cough tasks; 2) swallowing evaluation using videofluoroscopy; 3) voluntary cough acoustic recording; 4) completion of disease-specific rating scales including the ALS functional rating scale revised (ALSFRS-R)^41–42^; and 7) Quality of life/oral intake assessments including the Eating Assessment Tool (EAT-10)^43^, Functional Oral Intake Scale (FOIS)^44^.

### Swallowing Assessment

Safe vs. Unsafe swallowing was assessed using videofluoroscopy (VFSS). This testing methodology is a gold standard evaluation for swallowing function. Participants were seated in an upright position and a Phillips BV Endura fluoroscopic C-arm unit (GE OEC 8800 Digital Mobile C-Arm system type 718074) was used to collect radiographic images of the swallow in both a lateral and anterior-posterior viewing plane (30 frames/second). A Kay Pentax Swallowing Signals Lab unit (Kay Pentax, Lincoln Park, NJ) digitally recorded the fluoroscopic images that were stored for subsequent analysis. The standardized protocol consistent of the following bolus challenges: 1) 1-cc, 5-cc, 10-cc and cup sips of ultra-thin liquid contrast, 2) 5-cc of barium paste, and 3) ¼ of graham cracker coated with 5 cc barium paste in the lateral view; and 4) cup sips of ultra-thin barium liquid, 5) ¼ of graham cracker coated with 5 cc barium paste, and 6) 13-mm barium tablet in the anterior/posterior view. To ensure patient safety, the sequence of bolus presentations may have been altered. The VFSS study was discontinued if the patient silently aspirated and could not tolerate further swallow testing procedures.

Safe versus unsafe swallowing was determined using the standard protocol for swallow evaluation. The Penetration-Aspiration Scale score ^45^ was utilized to objectively assess swallowing safety, with scores of 1-2 classified as safe swallowing and scores of < 3-8 indicating unsafe swallowing with varying degrees of airway invasion. All assessments were evaluated clinically by the first author (SW), who analyzed the swallowing videofluoroscopy studies to identify the presence and extent of penetration or aspiration events according to the standardized scale criteria. Of the 23 motor speech disordered patients, 8 patients (3 PD; 5 ALS) were judged to have unsafe swallowing (PAS score > 3) as evaluated during any swallow task completed during the swallow study protocol.

### Acquisition of Acoustic Waveforms

Acoustic voluntary cough signals were collected for each participant (ALS, PD, and healthy controls). Patients were asked to “cough as if something were stuck in your throat.” An Audio-Technica microphone (ATM73a) was placed at a standardized distance of two inches from the patient’s oral cavity. Prior to collection of acoustic cough data, the acoustic signal was calibrated using a 90 dB pure tone. The microphone was routed to a preamplifier unit and the signal split into two A/D channels with each channel sampled at 40 kHz, 16-bits. Using software control, the sensitivity of each channel was set to allow simultaneous low and high-sensitivity recordings. This allowed for very high cough sound pressure levels to be recorded without clipping on the low-sensitivity channel and very low sound pressure levels to be recorded with an adequate signal to-noise ratio. Each subject was asked to produce a minimum of 3 trials with all recordings saved in .wav format. For the purposes of this study, the first cough from each participant was retained for a total of 33 cough samples (23 patients and 10 controls). The first cough in each epoch was selected for analysis to capture the pure, voluntary airway protective response to standard verbal stimuli before any conscious modulation or compensatory mechanisms could influence the acoustic signal. Figure 1 provides an example of the data collected for cough epochs in subjects with safe vs. unsafe swallows.

**Figure 1.**
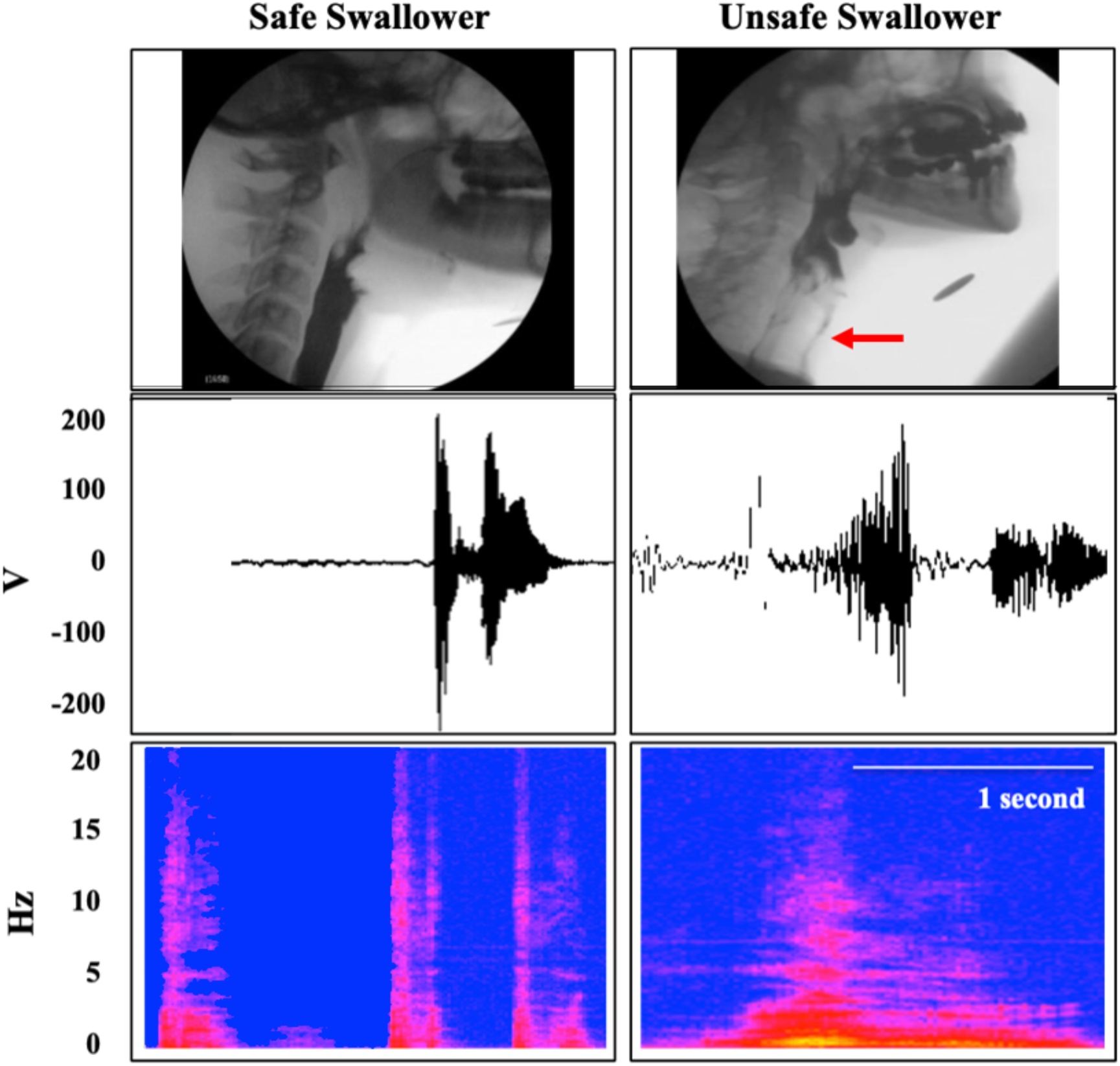
Voluntary cough profiles of ALS patients with safe (*PAS* ≤ 2) and unsafe (*PAS* > 2) swallowing. MBSS images of safe swallowing and unsafe swallowing (*top*) voluntary acoustic cough signals (*middle*) and corresponding spectrograms (*bottom*) are provided.

### Acoustic Analysis of Cough Signals

Before extracting features, cough audio files were edited using Audacity software tool to remove silence and extract relevant cough segments. Python 3.11.4 and the audio signal processing library Librosa 0.10.0 were used for extracting Mel Frequency Cepstrum Coefficients (MFCCs). Figure 2 provides a schematic of the MFCC analysis procedure. The edited cough segments were framed and windowed (frame length = 2048 points, corresponding to approximately 51 milliseconds with 75% frame overlap). Each frame was tapered using a Hanning window to minimize spectral leakage and introduction of spectral artifacts. The fast Fourier transform (FFT) spectrum was calculated for each frame, and the mel-frequency cepstrum was computed by applying mel-frequency filter banks to the logarithm of the FFT of the cough signal. Finally, the log-energy at the output of each filter is computed, with the MFCCs calculated via the discrete cosine transform (DCT) of the filter energy outputs. For each frame, 40 bands were calculated for the Mel spectrogram, with the first 13 MFCCs retained for analyses. While a large number of MFCCs will provide an extremely detailed representation of the spectrum, increasing numbers of coefficients represent very rapid changes in the spectrum that may be associated with extraneous noise and, therefore, may result in spurious information for classification. Thirteen MFCC coefficients have been used to effectively summarize the general shape and energy of the spectrum^36^ and have been successfully used in many applications including speech recognition^46–47^ identification of depression^31^, classification of disordered voice and resonance^48–49, 36,32,50,28^, and cough classification^25,51–52^. Using the aforementioned framing and windowing settings, a total of 1784 frames were extracted for the 33 cough samples. Given the analysis resulted in multiple frames per participant, the features were averaged across frames (mean number of frames = 54.3); there were no significant differences in the number of frames per group. The dataset was exported to comma-separated values (CSV) file for later analysis.

**Figure 2.**
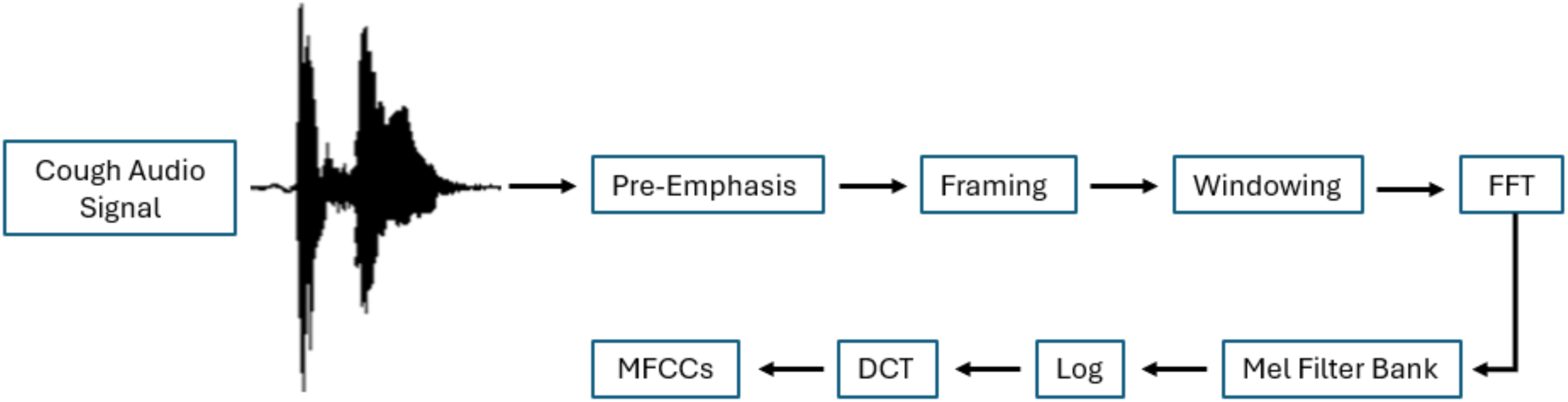
Schematic of the MFCC analysis procedure.

In addition to MFCCs, measures of zero crossing rate (ZCR), spectral centroid, spectral bandwidth, spectral roll off, and spectral contrast calculated for seven bands (0-200 Hz, 200-400 Hz, 400-800 Hz, 800-1600 Hz, 1600-3200 Hz, and 6400-12800 Hz) were also calculated for each data frame and averaged across frames per cough sample. This data was obtained to assess whether other acoustic measures could adequately classify subject grouping vs. MFCCs, as well as aid in the subsequent interpretation of individual MFCCs. The spectral centroid characterizes the center of mass of the spectrum, while the spectral bandwidth measures the spread or the variance of energy around the centroid. Spectral roll off is the frequency below which 85% of the spectral energy lies. ZCR is used to detect the presence voicing activity and is a measure of how many times the sign of the audio signal changes in the time domain. usually higher for unvoiced/aperiodic regions of the sound wave vs. lower for voice/periodic regions. Spectral contrasts were estimated by comparing the peak spectral energy to the valley spectral energy for each sub band.

### Statistical Analysis

The following comparisons were assessed; Healthy vs. Disordered (ALS+PD); Healthy vs. PD; Healthy vs. ALS, ALS vs. PD; and Safe vs. Unsafe swallow. For each of the aforementioned group comparisons, nonparametric Mann-Whitney U comparison tests were used to assess whether there were significant differences on the following acoustic variables: MFCCs (1-13); Spectral Centroid, Bandwidth, and Roll-off; Spectral Contrasts; and ZCR (converted to ms). Post-hoc effect sizes for Mann Whitney U tests were evaluated using biserial correlations. Spearman’s rho correlations were computed to ascertain the presence and strength of relationships between significant MFCC and the various specific spectral variables. Correlation coefficients range from 0 to ±1, with values from ±.1 to .3 considered weak; ±.4 to .7, moderate; and ±.8 to .99, strong to very strong^53^. As an exploratory study, no adjustments for family wise error were used in this analysis and effect sizes were used to ascertain the clinical significance of statistical findings.

Many studies extract a large number of features and classify with complex deep learning or machine learning models derived from very large samples. However, the use of a large feature set with relatively small clinical data sets as in the current study may increase the risk of overfitting^20^. Therefore, we elected to conduct ROC analysis for each comparison using the acoustic variable with the strongest group difference as assessed via the Mann Whitney U results. The ROC curve is created by plotting a point for each cutoff point score on the selected acoustic variable that represents the true positive score (i.e., sensitivity) on the ordinate and the false positive score (i.e., 1–specificity) on the abscissa. The performance of each ROC analysis was evaluated using area under the curve (AUC), sensitivity, specificity, overall model quality via the Gini Index (Gini Index – 2 x AUC -1). The ability of an acoustic variable to discriminate between comparative cases was represented by the area under the ROC curve (AUC). An AUC=1.0 is found for measures that perfectly distinguish between grouped individuals (e.g., healthy vs. disordered individuals). An AUC = 0.5 corresponds with chance-level diagnostic accuracy. In general, AUCs of 0.5–0.6 are considered almost useless; 0.6–0.7 poor; 0.7 to 0.8 fair; 0.8–0.9 good; and 0.9–1.0 excellent. Finally, various alternative cutoff scores were determined by identifying (1) cut-offs that maintained a best balance between sensitivity and specificity, and (2) cutoffs using the Youden Index (Youden Index = sensitivity + specificity-1), used as an alternative indicator of the optimal ROC cutoff point, with the maximum value of the index being 1 (when the test is perfect), and the minimum value being 0 (when the test has no diagnostic value).

## Results

### Healthy vs. Disordered Subjects (PD + ALS)

A series of initial Mann Whitney U tests indicated that healthy vs. disordered subjects differed significantly on MFCC 5, MFCC 6 with moderate effect sizes (see Table 3). The diagnostic accuracy and the precision of MFCC 5 (the acoustic variable with the strongest significance level) as a possible screening tool (i.e., to what extent MFCC 5 can distinguish between controls vs. disordered cases) was further examined using ROC analysis. Results indicated an AUC = 0.80 with Youden’s Index Se = 0.83 and Sp = 0.73 and a Balanced Se = 0.70 and Sp = 0.70. Table 4 provides ROC results and cutoff scores associated with Youden’s Index and balanced Se vs. Sp. Scores greater than the cutoff provide higher probability of a disordered cough signal vs a healthy cough. Figure 3 provides the ROC plot for the healthy vs. disordered cases.

**Figure 3.**
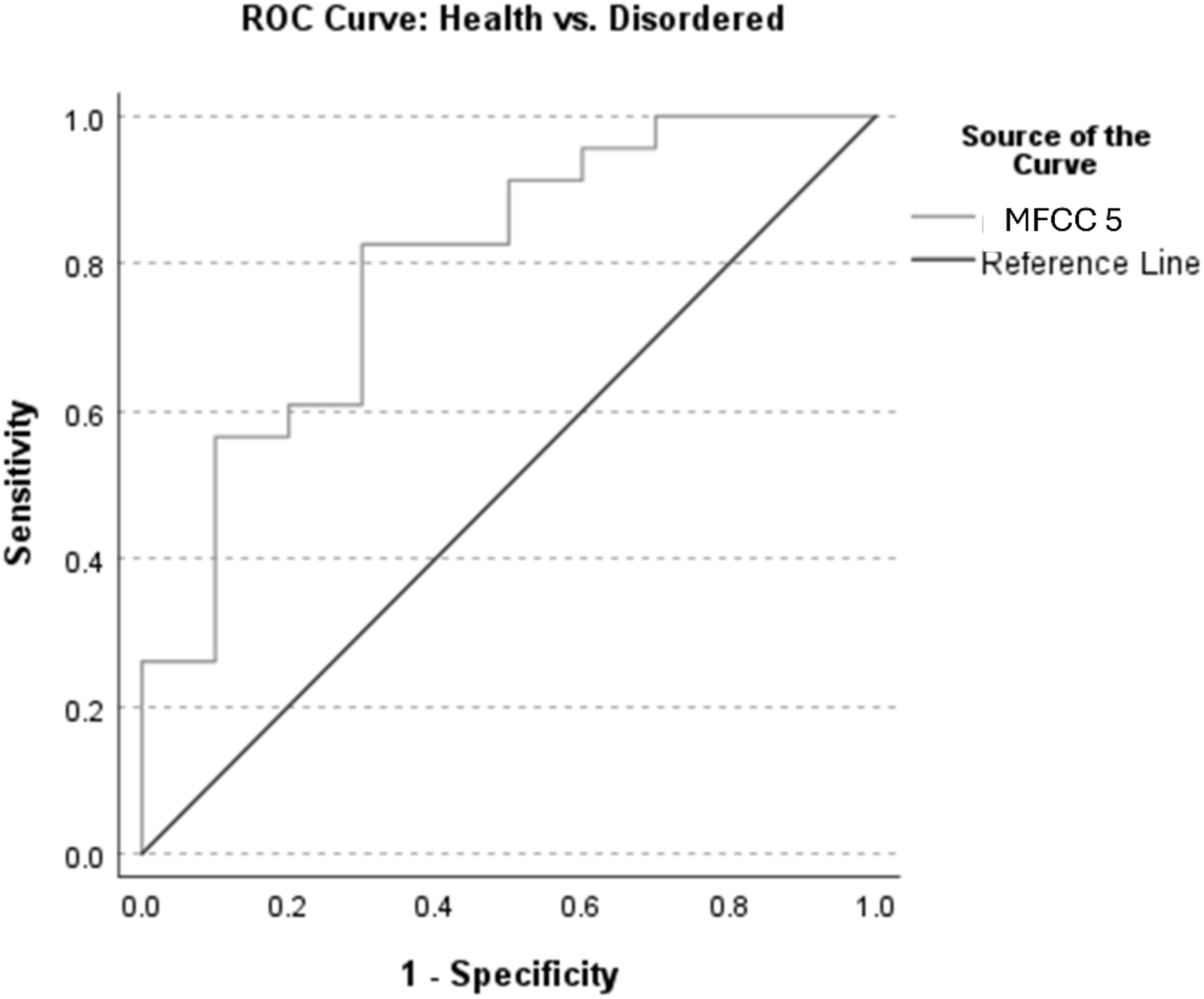
ROC curve for healthy vs. disordered (ALS + PD) cases. The AUC = 0.80.

**Table 3.**
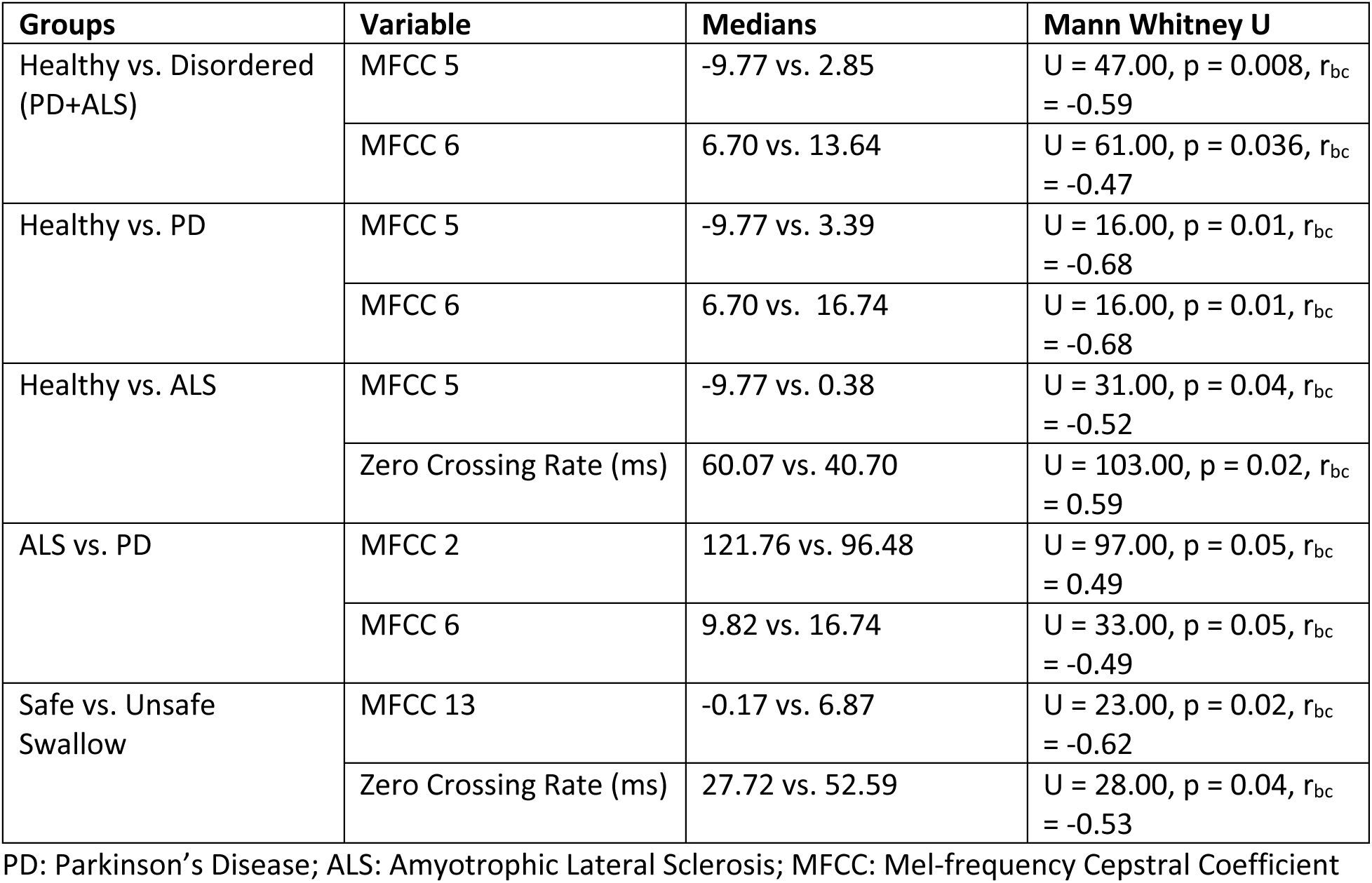
Table represents medians and Mann Whitney U scores across comparison groups.

**Table 4.**
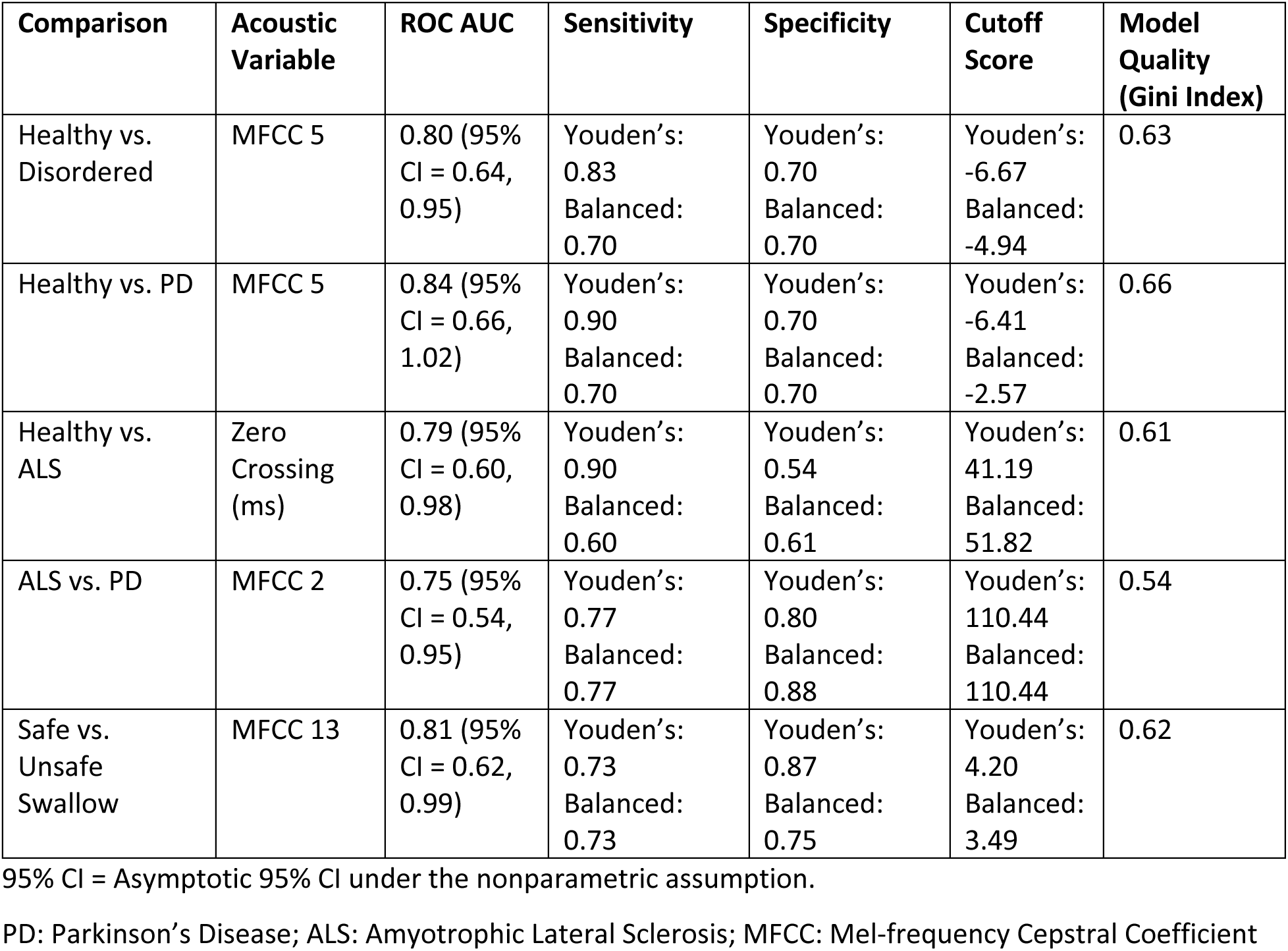
Comparison groups by variable with adjusted/balanced sensitivity and specificity measures using Youden’s J statistic.

| Comparison | Acoustic Variable | ROC AUC | Sensitivity | Specificity | Cutoff Score | Model Quality (Gini Index) |
| --- | --- | --- | --- | --- | --- | --- |
| Healthy vs. Disordered | MFCC 5 | 0.80 (95% CI = 0.64, 0.95) | Youden's: 0.83<br>Balanced: 0.70 | Youden's: 0.70<br>Balanced: 0.70 | Youden's: -6.67<br>Balanced: -4.94 | 0.63 |
| Healthy vs. PD | MFCC 5 | 0.84 (95% CI = 0.66, 1.02) | Youden's: 0.90<br>Balanced: 0.70 | Youden's: 0.70<br>Balanced: 0.70 | Youden's: -6.41<br>Balanced: -2.57 | 0.66 |
| Healthy vs. ALS | Zero Crossing (ms) | 0.79 (95% CI = 0.60, 0.98) | Youden's: 0.90<br>Balanced: 0.60 | Youden's: 0.54<br>Balanced: 0.61 | Youden's: 41.19<br>Balanced: 51.82 | 0.61 |
| ALS vs. PD | MFCC 2 | 0.75 (95% CI = 0.54, 0.95) | Youden's: 0.77<br>Balanced: 0.77 | Youden's: 0.80<br>Balanced: 0.88 | Youden's: 110.44<br>Balanced: 110.44 | 0.54 |
| Safe vs. Unsafe Swallow | MFCC 13 | 0.81 (95% CI = 0.62, 0.99) | Youden's: 0.73<br>Balanced: 0.73 | Youden's: 0.87<br>Balanced: 0.75 | Youden's: 4.20<br>Balanced: 3.49 | 0.62 |

95% CI = Asymptotic 95% CI under the nonparametric assumption.
PD: Parkinson's Disease; ALS: Amyotrophic Lateral Sclerosis; MFCC: Mel-frequency Cepstral Coefficient

MFCC 5 was observed to correlate strongly and significantly with spectral contrast (6400 to 12800 Hz) (rho = -0.84, p < 0.001), and moderately with Spectral bandwidth (rho = 0.56, p < 0.001), spectral contrast (800-1600 Hz) (rho = 0.58, p < 0.001).

### Healthy vs. Parkinson’s Disease (PD)

Mann Whitney U tests indicated that healthy vs. PD subjects differed significantly on MFCC 5 and MFCC 6 with moderate effect sizes (see Table 3). While the Mann Whitney U tests for MFCC 5 and MFCC 6 were identical, subsequent ROC analyses indicated that the model quality was stronger for MFCC 5 vs. MFCC 6. The ROC analysis evaluating the diagnostic accuracy and the precision of MFCC 5 as a possible screening tool for healthy vs. PD cases indicated an AUC = 0.84 with Youden’s Index Se = 0.90 and Sp = 0.70 and a Balanced Se = 0.70 and Sp = 0.70 (see Table 4). Scores greater than the cutoff provide higher probability of a PD cough signal vs a healthy cough. Figure 4 provides the ROC plot for the healthy vs. PD cases.

**Figure 4.**
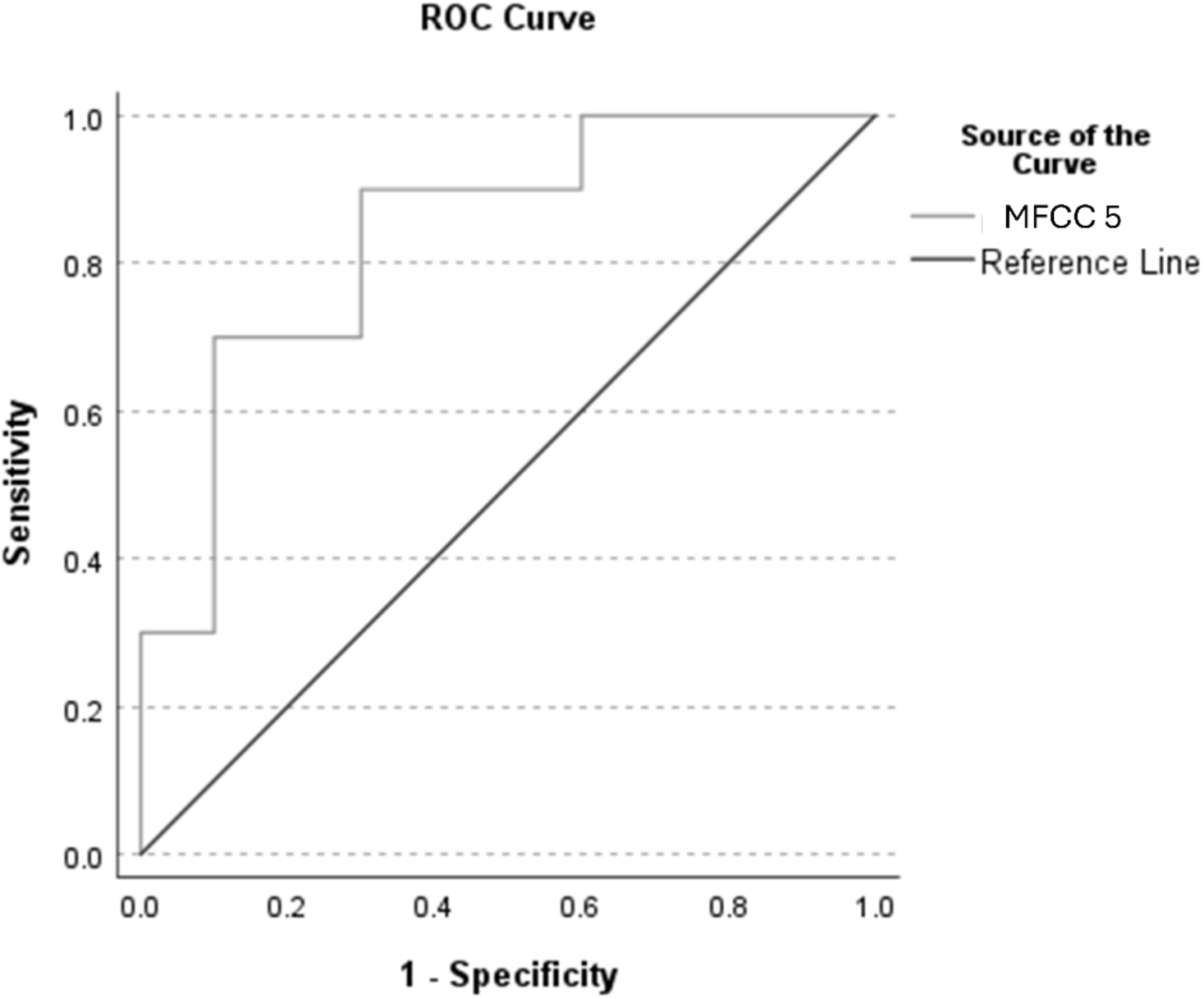
ROC curve for healthy vs. PD cases. The AUC = 0.84.

### Healthy vs. Amyotrophic Lateral Sclerosis (ALS)

Mann Whitney U tests indicated that healthy vs. ALS subjects differed significantly on MFCC 5 and Zero Crossing rate (ms) with moderate effect sizes (see Table 3). ROC using Zero crossing rate (ms) (observed as the acoustic variable with the strongest significance level) indicated an AUC = 0.79 with Youden’s Index Se = 0.90 and Sp = 0.54 and a Balanced Se = 0.60 and Sp = 0.61. In this case, scores greater than the cutoff provide higher probability of a healthy vs. a cough signal from an ALS participant. Figure 5 provides the ROC plot for the healthy vs. ALS cases.

**Figure 5.**
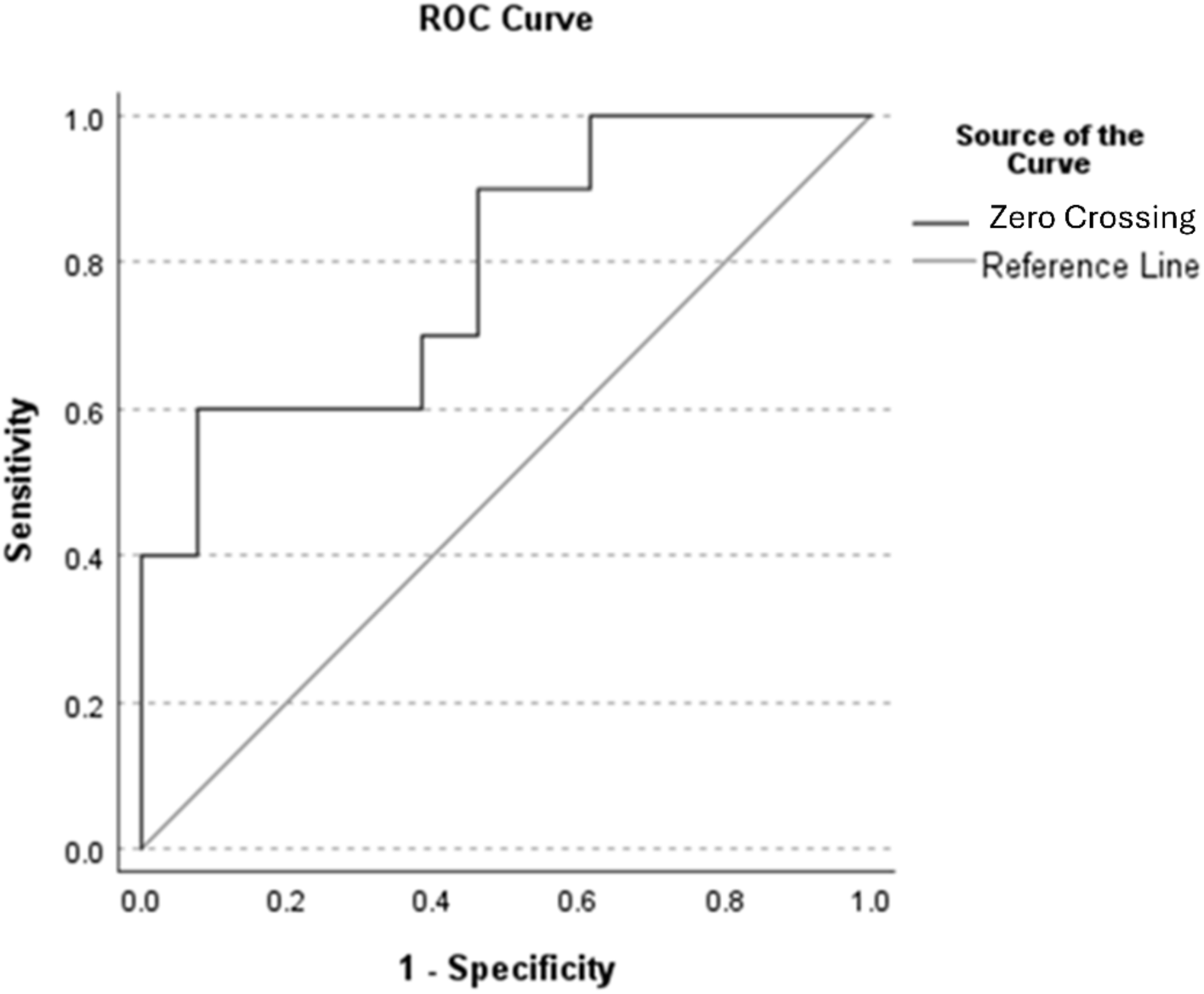
ROC curve for healthy vs. ALS cases. The AUC = 0.79.

### ALS vs. PD

Means comparison tests indicated that ALS vs. PD subjects differed significantly on MFCC 2 and MFCC 6 with moderate effect sizes (see Table 3). Mann Whitney U tests for MFCC 2 and MFCC 6 were identical though subsequent ROC analyses indicated that sensitivity and specificity results were stronger for MFCC 2 vs. MFCC 6. ROC using MFCC 2 indicated an AUC = 0.79 with Se = 0.77 and Sp = 0.80 (same results for both Youden’s Index and Balanced Se Sp). In this case, scores greater than the cutoff provide higher probability of a cough signal from an ALS vs. a PD participant. Figure 6 provides the ROC plot for the ALS vs. PD cases. MFCC 2 was observed to correlate strongly and significantly with the spectral centroid (rho = -0.84, p < 0.001), spectral bandwidth (rho = -0.78, p < 0.001), and spectral rolloff (rho = - 0.85, p < 0.001).

**Figure 6.**
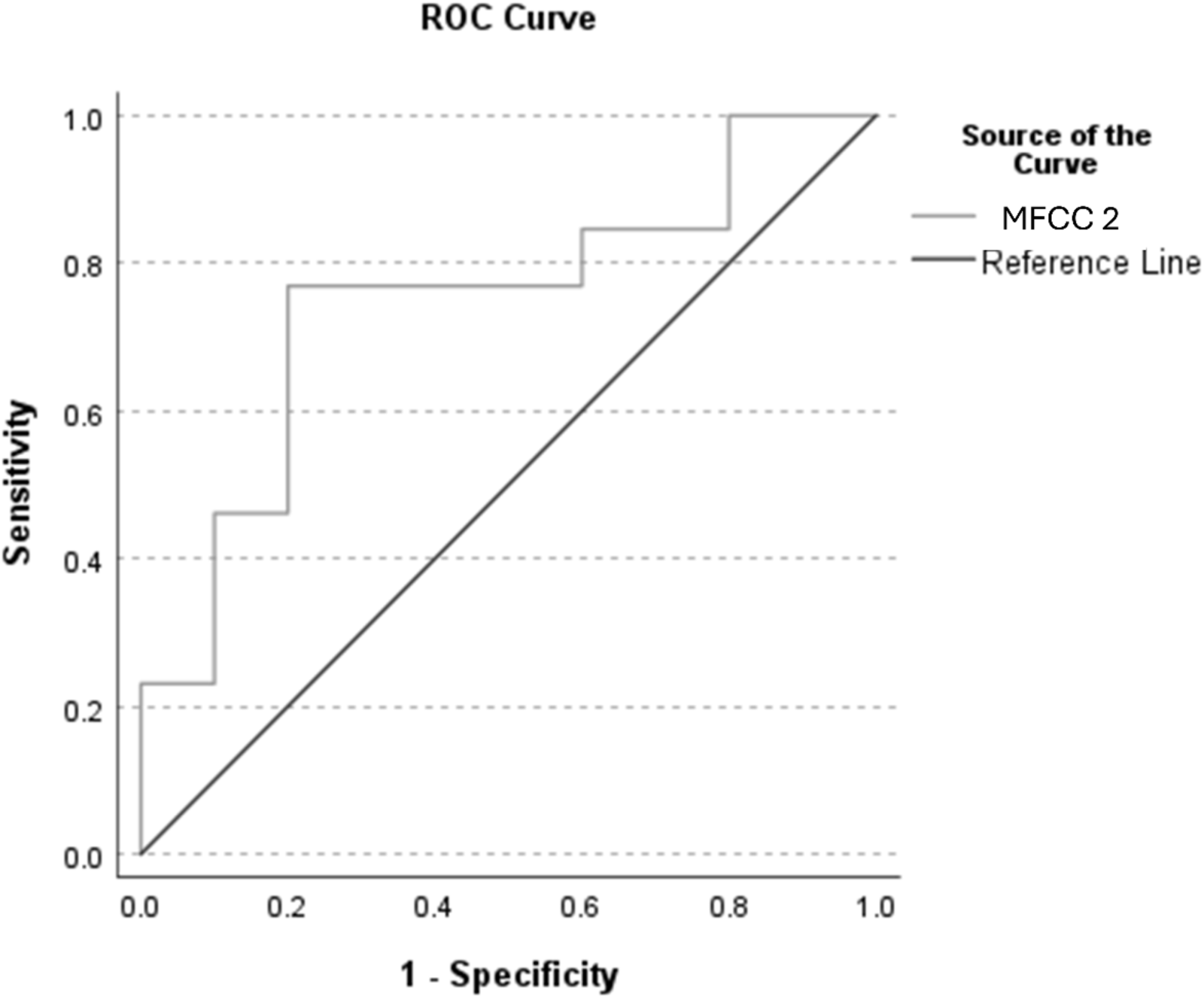
ROC curve for cases with ALS vs. PD cases. The AUC = 0.79.

### Safe vs. Unsafe Swallow

Means comparison tests indicated that subjects with safe vs. unsafe swallows differed significantly on MFCC 13 and zero crossing rate (ms) with moderate effect sizes (see Table 3). ROC using MFCC 13 indicated an AUC = 0.81 with Youden’s Index Se = 0.73 and Sp = 0.87 and a Balanced Se = 0.73 and Sp = 0.75. In this case, scores greater than the cutoff provide higher probability of a safe swallow vs. and unsafe swallow. Figure 7 provides the ROC plot for the ALS vs. PD cases. MFCC 13 was observed to correlate significantly with spectral contrast (1600-3200 Hz) (rho = -0.63, p = 0.001).

**Figure 7.**
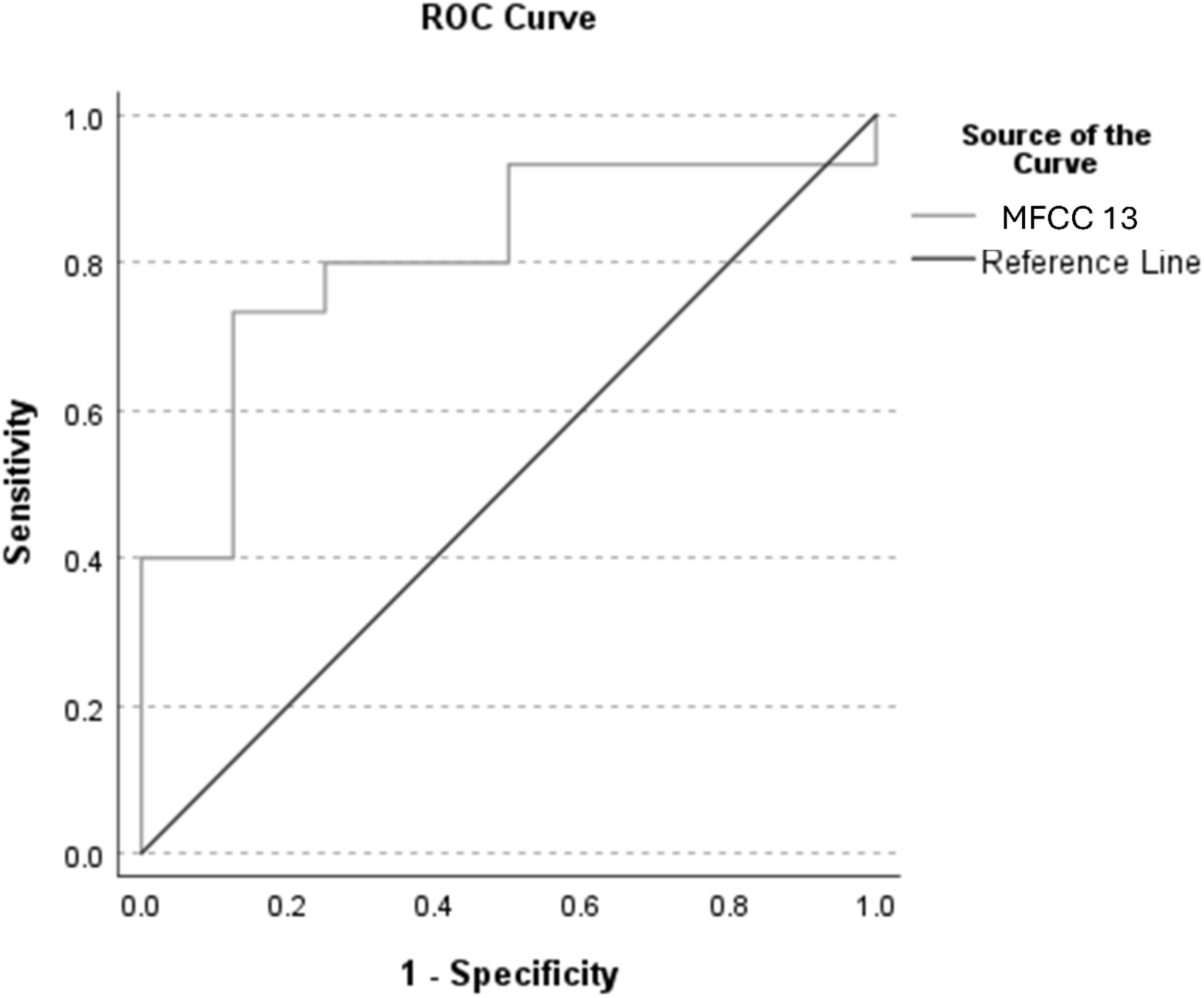
ROC curve for cases with safe vs. unsafe swallows. The AUC = 0.81.

## Discussion

MFCCs provide a representation of the overall shape of the spectral envelope information regarding the changes in its different spectrum bands. The results of this study confirm that cough creates a distinct sound pattern during the transition from a compression phase, or closure of the glottis, to expiration (Korpáš et al., 1996), and MFCC’s provide an effective and efficient means of detecting aberrant characteristics which may indicate disease state or swallowing safety status.

### Healthy vs. Motor Disordered Subjects

The results of this study show that a selected set of MFCC’s are able to effectively discriminate between healthy vs. motor disordered subjects. For both healthy vs. combined PD+ALS and healthy vs. PD subjects, MFCC5 was observed as the strongest discriminator with an ROC area under the curve (AUC) ≥ 0.80 (see Table 4). The AUC is a widely reported measure of the accuracy of a diagnostic test and indicates the ability of a test or measure to discriminate between cases and non-cases/controls.

These results indicate that MFCC 5 is a fair-to-good discriminator with higher values for this coefficient (i.e., greater than the cutoff) generally indicative of a disordered cough. MFCC 5 was also observed to have a significant and strong inverse correlation the spectral frequency band between 6400-12800 Hz, with greater MFCC 5 values associated with decreased energy in this contrast band. In addition, the positive value for MFCC 5 for disordered subjects vs. the negative MFCC for healthy has been said to be indicative of spectral energy concentrated in lower frequency regions vs. higher frequency spectral energy with negative MFCC’s^36, 55^. While these acoustic measures are indirect measures of underlying physiology, it may be speculated that weaker explosive/expulsive energy in the initial cough phase may result in reduced higher frequency spectral energy in the 6400-12800 Hz band. In addition, the negative MFCC 5 value for healthy subjects vs. disordered is again indicative of an increase in high frequency spectral energy during the cough.

While MFCC 5 was also a strong discriminator between healthy vs. ALS, the zero crossing rate (ZCR) emerged as a slightly stronger factor. ZCR is commonly used as an indicator of periodicity in the acoustic signal, with lower values associated with more periodic activity (fewer zero crossings per unit time) vs. higher values associated with higher frequency, more aperiodic activity. In this study, healthy subjects were observed to have higher ZCR vs. ALS and may again indicate the presence of higher frequency explosive/expulsive acoustic activity in the healthy cough. Recent work in acoustic analysis by Rocha et al. (2024)^56^ explored relationships between cough sounds and characteristics from respiratory and bulbar domains in ALS. They reported significant differences in spectral frequency characteristics (spectral bandwidth, spectral centroid, spectral roll-off, spectral kurtosis, and spectral skewness) between ALS patients and healthy controls. The results of this study also show that spectral features captured via MFCCs, as well as other acoustic features (such as ZCR) of voluntary cough signals, can differentiate between health states, neurological disease types, and swallowing safety status.

### PD vs. ALS

The ability to effectively categorize the cough signals of healthy vs. motor speech disordered subjects is certainly clinically beneficial. However, a more difficult and perhaps even more beneficial prospect is the ability to successfully categorize disordered subjects who have a related underlying pathological signs and symptoms. Though ALS is categorized as a mixed spastic-flaccid dysarthria vs. the hypokinetic dysarthria of PD, both types of dysarthria may be characterized by slowness in movement, weakness, and limited range of motion. While there are similarities in motor disruption between these two disorders, the results of this study showed that MFCC 2 was an effective discriminator of ALS vs. PD with an observed AUC = 0.79 and respectable Se = 0.77 and Sp = 0.80. MFCC2 has been interpreted as a generalized low-to-high spectral energy ratio (Tracey et al., 2023), representative of broad spectral features and relatively slow variations in the spectral envelope. This interpretation of MFCC 2 is supported by the observation that, in this data set, MFCC 2 was observed to correlate strongly and significantly with general spectral characteristics such as the spectral centroid (rho = -0.84, p < 0.001), spectral bandwidth (rho = -0.78, p < 0.001), and spectral roll off (rho = -0.85, p < 0.001). A greater MFCC 2 value was observed for ALS (see Table 3), indicating a greater low vs. high spectral ratio for ALS vs. PD.

Many PD patients have often been characterized as having asthenic and breathy voice production with possible vocal fold bowing^57–58^. If present, these characteristics may also be expected to potentially affect the voiced Phase 3 of the typically described cough signal, and the presence of a high frequency spectral emphasis (i.e., reduced low vs. high spectral ratio) may be expected in the PD cough. It is likely that insidious motor dysfunction across the neural axis contributed to these findings, as lesions anywhere along the neuraxis may impact cough motor and subsequently acoustic function. Physiological impairment leading to dystussia may originate from airway afferent impairment, respiratory muscle weakness, or a combination of both. In terms of motor impairment, chest wall rigidity in patients with PD decreases their ability to inflate the lungs^3^. The work of Ebihara and colleagues (2003)^11^ indicated that involvement of motor and sensory impairment might depend on the stage of PD progression. In contrast, the ALS patient population presents in a heterogeneous manner, making evaluation and benchmarking of impairment difficult using clinical indices such as perception via rating scales as well as with objective acoustic measures.

### Safe vs. Unsafe Swallow

There are relationships between cough and swallow at both neural^59–61^, and anatomical/functional levels^62,3,5^. ALS simultaneously results in motor cough deficits,^5^ and sensory cough deficits^63^. Similarly, both motor and sensory components of cough may be impaired in PD due to factors such as cough reflex impairment and chest wall rigidity ^11–12^. Within the motor speech disordered subjects (i.e., PD and ALS subjects combined), MFCC 13 was observed as the most effective variable in separating the cough signals of subjects with safe vs. unsafe swallowing subjects. Though higher-order MFCCs have been said to capture faster variation across the frequency spectrum often representative of noise or fine details (Tracey et al., 2023), MFCC 13 was observed in the current study to be moderately inversely correlated (rho = -0.63) with spectral contrast in the mid-frequency 1600-3200 Hz band (i.e., in this study, an increased MFCC 13 value was associated with a decrease in energy in the 1600-3200 Hz frequency band). As previously mentioned, the sign of a Mel-Frequency Cepstral Coefficient (MFCC) may also provide insight into the distribution of spectral energy across frequencies in a sound signal. The cough of safe swallowing subjects was observed to have a negative sign which suggests that most of the spectral energy is concentrated at high frequencies and may coincide with the greater energy in the 1600-3200 Hz band observed for safe swallowing subjects. In contrast, the positive value for MFCC 13 observed for the unsafe swallowing subjects may indicate that the majority of the spectral energy is concentrated in the low-frequency region of the frequency band of interest (1600-3200 Hz). This speculation is consistent with Figure 1 which shows examples of ALS patients with safe vs. unsafe swallows. In this example, the unsafe swallow is observed to have more spectral energy in the lower region of the spectrogram and contrasts with the safe swallow example in which broadband energy extending into the 15-20 kHz region is observed during the explosive phases of the cough.

### Limitations

The findings reported in this study are limited by several factors. Acoustic analyses were computed across all phases in the first cough bout of a cough episode. Future studies may focus analyses of acoustic characteristics in coughs signals in alternative cough bouts as well as in individual cough phases to further determine the locus of differences in cough signals obtained from healthy vs. disordered subjects. Because acoustic analyses provide indirect measures of the cough signal, future studies that obtain simultaneous acoustic, physiologic, and imaging data may be able to provide greater insight and/or confirmation as to the underlying differences in cough physiology that may lead to acoustic differences between heathy vs. disordered or within disordered differences in the cough signal. An additional limitation is that this study only used one rater for assessment of swallowing safety status (primary author) without reliability analysis. Future work may include a more robust sample of patient coughs, a more diverse set of patient disease states and multiple raters to assess the perceptual aspects of the recorded cough signal.

### Conclusions

Cough creates a distinct sound pattern during the transition from a compression phase, or closure of the glottis, to expiration. The audible and audio recordable nature of cough provides an opportunity to detect aberrant characteristics which may indicate disease state or swallowing safety status and, unlike traditional perceptual assessments of cough which can be unreliable, these objective acoustic measures offer readily available quantifiable parameters that capture complex physiological impairments across the neural axis affecting both cough and swallow functions. Our findings demonstrate that, even with a conservative approach to acoustic analysis of voluntary cough signals, MFCC analyses, along with other acoustic features including ZCR, show strong potential for distinguishing between disease states and swallowing safety status. The relevance of such work is paramount for professionals who serve populations with impaired swallow ability, as dystussia promotes poor airway protection and must be a target of evaluation and intervention. Future research should focus on expanding these preliminary findings with larger cohorts and developing standardized acoustic analysis protocols that can supplement clinical evaluations, potentially serving as early biomarkers for disease progression and swallowing safety in neurogenic populations.

## Data Availability

All data produced in the present study are available upon reasonable request to the authors.

## Acknowledgments

The first author would like to acknowledge the support of doctoral mentors Drs. Emily Plowman, David Eddins, and Kendall Morris for ongoing support and contribution of resources for the collection of data.

